# Impact of direct-acting antiviral treatment of hepatitis C on the quality of life of adults in Ukraine

**DOI:** 10.1101/2021.09.08.21263288

**Authors:** M Benade, S Rosen, S Antoniak, C Chasela, Y Stopolianska, T Barnard, MM Gandhi, I Ivanchuk, V Tretiakov, J Dible, T Minior, KW Chew, C van der Horst, Z Tsenilova, I Sanne, for EQUIP Health

## Abstract

**Background:** Direct-acting antivirals (DAAs) are highly effective in achieving sustained virologic response among those with chronic hepatitis C virus (HCV) infection. Quality of life (QOL) benefits for an HCV-infected population with high numbers of people who inject drugs and people living with HIV (PLHIV) in Eastern Europe have not been explored. We estimated such benefits for Ukraine.

**Methods:** Using data from a demonstration study of 12-week DAA conducted in Kyiv, we compared self-reported QOL as captured with the MOS-SF20 at study entry and 12 weeks after treatment completion (week 24). We calculated domain scores for health perception, physical, role and social functioning, mental health and pain to at entry and week 24, stratified by HIV status.

**Results:** Among the 857 patients included for the final analysis, health perception was the domain that showed the largest change, with an improvement of 85.7% between entry and week 24. The improvement was larger among those who were HIV negative (104.4%) compared to those living with HIV (69.9%). Other domains that showed significant and meaningful improvements were physical functioning, which improved from 80.5 (95% CI: 78.9-82.1) at study entry to 89.4 (88.1-90.7) at 24 weeks, role functioning (64.5 [62.3-66.8] to 86.5 [84.9-88.2], social functioning (74.2 [72.1-76.2] to 84.8 [83.2-86.5] and bodily pain (70.1 [68.2-72.0] to 89.8 [88.5-91.1]). Across all domains, QOL improvements among PLHIV were more modest than among HIV-negative participants.

**Conclusion:** QOL improved substantially across all domains between study entry and week 24. Changes over the study period were smaller among PLHIV.

## Background

Viral hepatitis C infection (HCV) remains a major public health concern globally, with 71 million people estimated to be living with chronic HCV infection and at high risk of developing liver cirrhosis, hepatocellular carcinoma, liver failure, and death if left untreated.^1–3^ DAA treatment, which cures HCV within 8-12 weeks, is now widely available in some countries.^4^ Benefits in treating HCV are found in averted deaths, reduced morbidity, and improvement of reported QOL and economic productivity.^5–8^

In Ukraine, it is estimated that 3.5% of the general population are living with HCV, of whom a large proportion are people who inject drugs (PWID) and many are co-infected with HIV.^9^ Other high risk groups include sexual partners of those with HIV, sex workers, prisoners, military members, and populations in conflict zones.^9^ Lack of adequate state funding, especially for diagnostics, presents a significant barrier to accessing treatment with DAA.^10^

In 2018, a single-arm demonstration project was implemented to evaluate a scalable, integrated, low cost HIV and HCV testing and management intervention in Ukraine offering HCV treatment with sofosbuvir/ledipasvir for 12 weeks, with or without weight-based ribavirin (WBR) in Ukraine.^11^ The study also collected data on participants’ self-reported QOL at the start and end of the study period, which we present here.

## Methods

### Parent study

For the original demonstration study, patients were enrolled at two public clinics in Kyiv, Ukraine. Study sites, sample selection and data collection have been described previously.^11^ Eligibility criteria included HCV viraemia, being treatment naïve and age ≥18 years, and no or compensated liver cirrhosis. Those eligible were offered HIV and HCV testing, 12-week treatment of HCV with DAAs supported by simplified HCV treatment monitoring, and initiation of HIV antiretroviral therapy (ART) for those with untreated HIV co-infection. The study’s primary outcome was sustained virologic response (SVR) at 12 weeks post treatment completion (24 weeks). Of the 868 participants enrolled, 860 completed the 24-week follow-up visit and were assessed for SVR. By intention-to-treat analysis, the overall treatment success rate was 95.7% (831/868).^11^

### Quality of life data and analysis

Interviews were conducted at study entry and 24 weeks for QOL indicators, using an adapted version of the Medical Outcomes Study 20-Item Short Form Health survey (MOS SF-20) (supplementary file 1).^12^ We compared QOL measures between baseline and endline for each question. We also aggregated responses according to MOS SF-20 categories of health perception, physical functioning, mental health, role functioning, social functioning, and bodily pain to assign a mean value for each category at baseline and endline, stratified by HIV status. Scores for each of the six subscales (physical, social and role functioning, pain, mental health, and health perception) were calculated as described by Stewart et al^12^. In each domain, the lowest possible score was zero and the highest possible score was 100. We stratified the domain scores by HIV status and performed t-tests to determine statistical significance in difference of means between baseline and endline, as well as by HIV status and major demographic characteristics (age, education, employment status, injection drug use status).

## Results

Of the 868 patients who were included in the primary analysis, we assessed baseline data on all 868 and endline data on 857, with the discrepancy reflecting patients who did not complete the study or had substantial missing data on QOL variables. Detailed characteristics of enrolled participants, their clinical and laboratory outcomes, and the costs of the intervention to the provider are reported elsewhere^11^ and summarized in supplementary table 1. The median (IQR) age of the population was 39 years (35-44) and 66% were male. Most participants identified as PWID (87%), of whom 95% self-reported being on medication-assisted therapy (MAT). Just over half the sample (482, 55%) were HIV positive; all were on ART at the time of study enrollment. The 11 patients who were excluded from the final analysis did not differ from those in the analysis in terms of employment status or education level; all those excluded identified as PWID; were younger than 45 years, did not have liver cirrhosis, and were more likely to be co-infected with HIV; and 90% were male (supplementary table 2).

Responses to all QOL questions at baseline and endline are reported in supplementary table 3. Below we highlight differences between the two periods in the six domains listed above.

### Self-reported health

At study entry, 66% of participants reported their health as either “poor” or “fair”. By study exit, 80% reported their health as “good”, “very good” or “excellent.” Consistent with this result, 73% of participants reported that their health was either “somewhat better” (43%) or “much better (30%) at 24 weeks than at study entry. The proportion of patients who agreed with the statement “my health is excellent” increased from 27% to 68% over the study period, while those agreeing with the statement “I have been feeling bad lately” decreased from 57% to 11%. The mean domain score for health perception increased from 33.7 (95% CI 32.4-35.1) at study entry to 62.3 (60.7-63.9) at 24 weeks (Figure 1).

**Figure 1.**
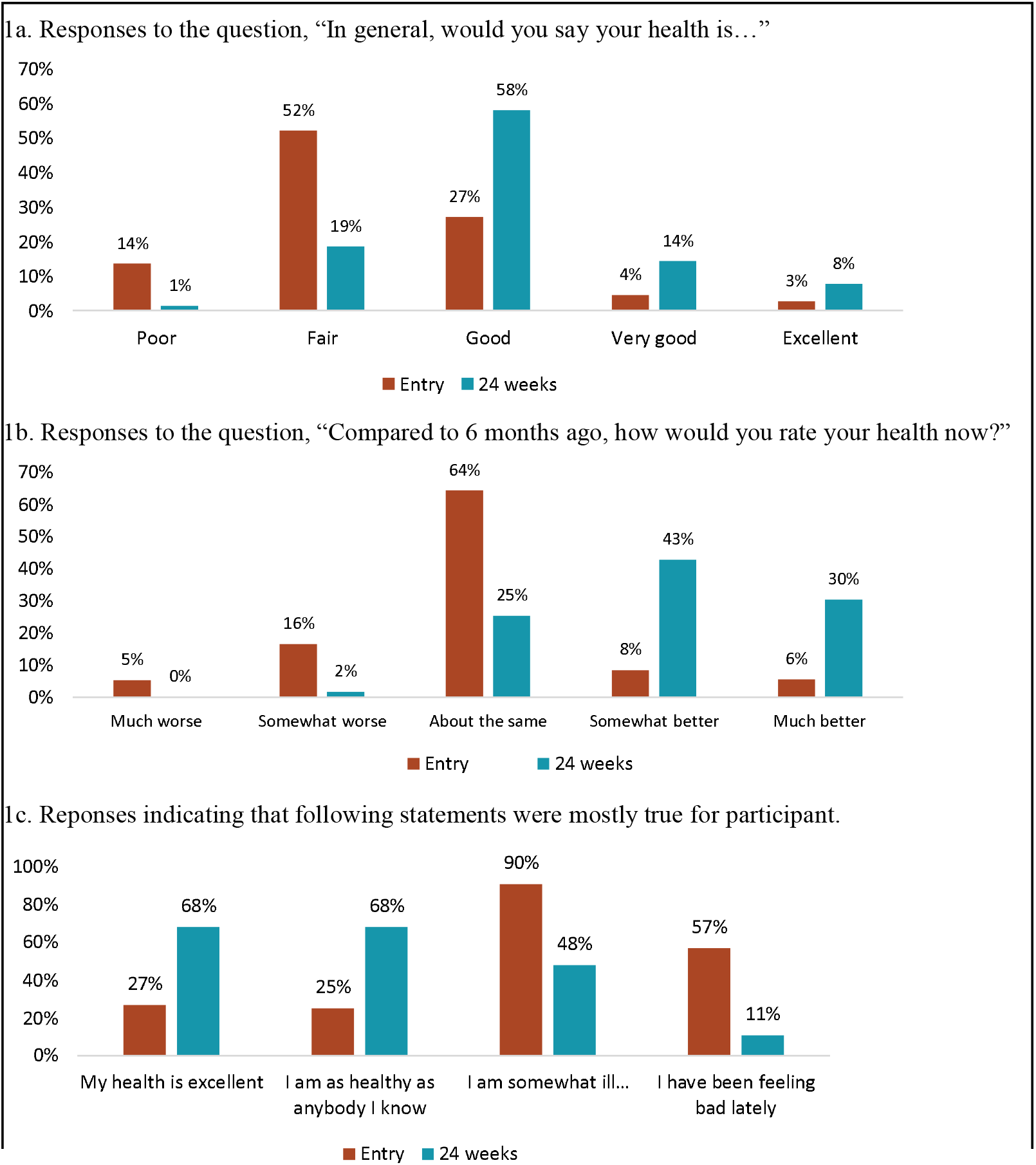
Self-reported health at study entry (baseline) and 24 weeks.

### Physical activities, role, and social functioning

The proportions of participants who reported being limited in conducting a range of physical activities fell over the study period for all activities (Figure 2). The largest difference was seen in vigorous activity, where the proportion who reported facing limitations declined from 54% to 32%. The mean domain score for physical functioning increased from 80.5 (95% CI 78.9-82.1) to 89.4 (88.1-90.7). Formal sector employment also rose over the study period, from 33% to 40% of the cohort. The proportion of participants who reported that their health affected “a lot” the quality at which they could perform their main activity decreased from 76% at study entry to 12% at 24 weeks. The mean number of days that participants were unable to perform their main activity due to their health or seeking health care in the past 2 weeks decreased from 1.6 (95% CI 1.5-1.8) days to 0.8 (0.6-1.0) days over the study period. Finally, the mean social functioning score improved from 74.2 (95% CI 72.1-84.8) to 84.8 (83.2-86.5).

**Figure 2.**
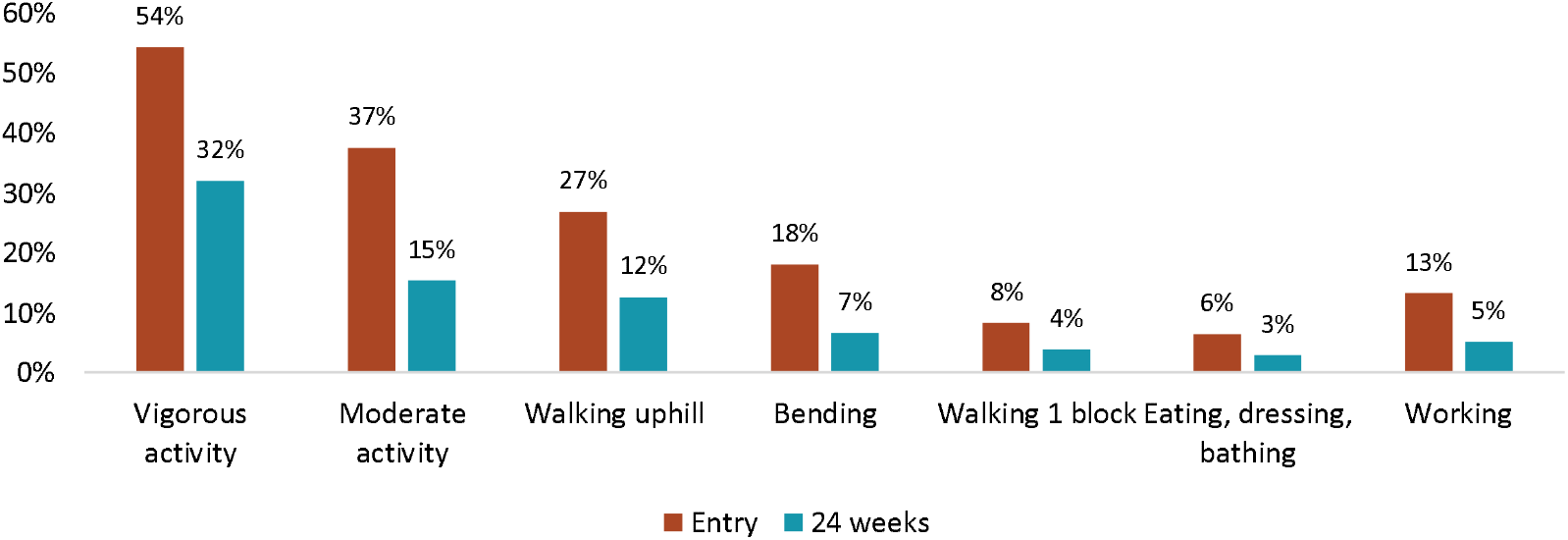
Functional impairment at study entry (baseline) and 24 weeks Response to the question, “My health limits me in doing the following activities…”

### Mental health and symptoms

Questions related to anxiety and depression showed a substantial improvement in mental health over the study period (Figure 2), with the mean score for the mental health domain increasing from 67.0 (95% CI 65.5-68.2) to 70.5 (69.1-66.9).

The proportion of patients who reported having no or very mild inability to focus increased from 54% at study entry to 77% at endline, while those who reported a moderate inability to focus decreased from 23% to 9%. An improvement in reported tiredness and fatigue was also observed, increasing from 21% who reported no or very mild fatigue at study entry to 62% at 24 weeks. Bodily pain improved from 50% reporting no or very mild body pain at initiation to 84% at study endline. The mean domain score for pain improved from 70.1 (95% CI 68.2-72.0) to 89.8 (88.5-91.1) over the study period.

### Stratification by HIV status

Participants with HIV and those without did not differ at baseline for any of the domains (supplementary table 3). Domain scores reported at 24 weeks in the physical functioning domain were lower for PLHIV (87.4, 95% CI 85.5-89.4) than for those not living with HIV (91.9, 90.0-93.5). Improvements in the social domain were larger among those who were not living with HIV (87.7, 95% CI 85.5-89.9) than among those with HIV (82.5, 80.1-84.9). Similarly, health perception improved by a larger margin among those who were not living with HIV (67.9, 95% CI 64.5-70.3) than among those with HIV (57.7, 55.6-59.9). Role functioning was slightly higher among those not living with HIV (87.2 (95% CI 84.7-89.8) vs 85.8 (83.5-88.1)) and domain scores for pain and mental health were marginally higher among PLHIV, but these differences were not statistically significant.

Other variables such as education, age, employment status and whether or not participant was considered PWID were also examined but there were no differences in these indicators between people with and without HIV.

## Discussion

In this study of more than 850 HCV-infected adults in Ukraine, of whom most were PWID and over half had HIV, effective HCV treatment produced large gains in QOL across all measured domains. The greatest improvements between study enrollment (baseline) and 24 weeks (endline) were in health perception (+26.8), role functioning (+22.0), and pain (+19.7). Social functioning (+10.9) and physical functioning (+8.9) improved somewhat less but still showed significant improvement at endline compared to study entry.

The results we observed in this cohort in Ukraine were similar to or of greater magnitude than those reported from other settings.^5,13–15^ In a Japanese cohort, the only domain of the MOS SF-36 that was found to be significantly improved was general health perception, with an improvement of 3.81 that persisted at 3 years after treatment with DAA.^5^ Younossi et al found similarly that statistically significant, modest improvements in reported QOL were sustained as long as 2 years after SVR was achieved in U.S. patients.^16^ The larger improvements we observed may reflect our relatively short follow up period (24 weeks after treatment initiation) and the fact that our study was conducted in a middle income country, rather than high income, and among many patients with co-morbidities; our participants had worse baseline domain scores and thus more room for HCV treatment to improve their QOL.

Our study had several limitations. Data were drawn from a single-arm implementation trial; we had no HCV-negative comparison population. Because the treatment success rate was so high, we also lack a comparison with those failing HCV treatment. It is possible that some of the improvements we observed would have come about anyway, without HCV treatment, though the relatively short interval between our baseline and endline interviews (24 weeks) makes major secular shifts unlikely. By definition, patients who were lost to follow up were missing endline data; if these patients were also sicker or more disabled by HCV than were those who did finish, our results would overestimate overall improvements. Comparison of the two groups at baseline, however, did not reveal any differences in their condition. Our choice of tool, the SF-20, is known to have floor effects for social functioning, bodily pain, and role functioning and ceiling effects across all domains reported. Finally, our endline measurement took place 12 weeks after treatment completion, and therefore findings are limited to short-term improvements in QOL only.

Despite these limitations, the large improvements we observed in QOL for this cohort of patients suggests that HCV treatment, as delivered in this intervention, has a major positive effect on the mental and physical well-being, self-perception, and productivity of those who complete treatment. While some of the impacts are slightly more modest for patients with HIV, general conclusions remain the same. These results add to the benefits of offering effective HCV treatment to all with active HCV infection.

## Data Availability

Data is available upon request

## Funding

This work was supported by USAID EQUIP Grant No. AID-OAA-A-1500070 to Right to Care and Luxemburg Business Partnership Facility 2017, Grant No. MAE/014 – 17 1620.

## Competing interests

KWC has received a research grant to the institution from Merck Sharpe & Dohme. The remaining authors declare that they have no competing interests.

## Authors’ contributions

IS, SR, CC, KWC, CVDH, TB, SA, conceived the study; SA, YS, TB, II conducted the study; MB, SR, CC, KWC, MMG analyzed the data; MB, SR drafted the manuscript; all authors reviewed and approved the final manuscript.

## Notes

### Competing Interest Statement

Dr Kara W. Chew has received a research grant to the institution from Merck Sharpe & Dohme. The remaining authors declare that they have no competing interests.

### Clinical Trial

NCT04038320

### Author Declarations

The study was reviewed by the Ukranian Institute on Public Health Policy IRB #1, Institutional Review Board (00007612) and the University of Witwatersrand Human Research Ethics Committee (M17078). The Boston University Institutional Review Board approved analysis of a de-identified analytic dataset (H-37820). All participants provided written informed consent.

